# Spatial and Temporal Patterns and Clustering of Routine Childhood Immunization Coverage across Districts of North-Western Province, Zambia, 2021–2025

**DOI:** 10.64898/2026.09.14.26363025

**Authors:** Marshall Chitalu Mubanga, Friday Nkoma, Eustarckio Kazonga, Martin Chakulya, Chipego Hajamba

## Abstract

**Background:** **Routine childhood immunization a cornerstone of child survival,** substantial subnational disparities persist in many low- and middle-income settings. Evidence on the spatial and temporal distribution of routine immunization coverage at district level in North-Western Province, Zambia, is limited**. This study examined** spatial and temporal patterns and clustering of routine childhood immunization coverage across the province’s 11 districts from 2021 to 2025.

**Methods:** A retrospective ecological study was conducted using annual district-level routine childhood immunization coverage data from North-Western Province, Zambia, for 2021–2025. Data were obtained from routine health information system records. Descriptive statistics were used toassess temporal and district-level variation.. Spatial inequality was quantified using the range, interquartile range (IQR), and coefficient of variation (CV). Global spatial autocorrelation was evaluated using Global Moran’s I, while local spatial patterns were quantified using Local Moran’s I (LISA) and Getis-Ord Gi* hotspot analysis. First-order Queen contiguity with row-standardized spatial weights: was used to define spatial relationships. Statistical significance was assessed using 999 permutations at α = 0.05.

**Results:** Coverage varied substantially across districts and years, ranging from 20.7 % (Chavuma, 2023) to 108.5 % (Kasempa, 2025). Spatial inequality as greatest in 2023 (CV 37.3 %) and declined to 18.1 % in 2025.Global spatial autocorrelation was weak and generally non-significant across most years.

Local analysis identified Chavuma as a statistically significant Low-High spatial outlier, while Kasempa and Mufumbwe formed a significant High-High cluster: in 2005. Getis-Ord Gi* similarly identified Kasempa and Mufumbwe as significant hotspots in 2025 and detected no significant cold spots during the study period. Mean district-level coverage declined by approximately 3 percentage points between 2021 and 2025, with Mean district-level coverage declined by approximately 3 percentage points between 2021 and 2025, with large declines in Solwezi (–47.9 pp) and Zambezi (–20.0 pp) while Kasempa (+26.1 pp) and Mufumbwe (+16.5 pp) recorded increases.

**Conclusions:** Immunization coverage in North-Western Province exhibited substantial temporal and district-level variation, with generally weak global spatial autocorrelation. However, a statistically significant high-coverage cluster emerged in 2025 in Kasempa and Mufumbwe. The transient Low-High outlier in Chavuma in 2023 further highlights the importance of monitoring geographically isolated areas of low coverage. Findings support spatially informed micro-planning, targeted outreach, and continued district-level surveillance to address persistent inequalities and emerging geographic patterns in immunization coverage.

## 1.0 Introduction

Childhood immunization is one of the most effective public health interventions for preventing morbidity and mortality from vaccine-preventable diseases (1). Global and regional data show that high coverage with basic vaccines such as diphtheria, tetanus, pertussis (DTP), measles-containing vaccine and newer antigens substantially reduces under-five mortality and contributes to progress towards child health targets. Despite overall gains, many low-and middle-income countries continue to experience pockets of low coverage, persistent zero-dose children and frequent missed opportunities for vaccination, especially in geographically remote and socio-economically disadvantaged communities (2,3).

Zambia has made sustained investments in its Expanded Programme on Immunization, achieving relatively high national coverage for key antigens, including pentavalent vaccine and measles-containing vaccine, across multiple data sources. However, survey data and provincial analyses highlight that full immunization coverage among children remains suboptimal and unevenly distributed (4). For example, national survey results have reported that roughly three-quarters of children aged 12–23 months have received all basic vaccinations, with coverage varying by region, maternal education and other socio-demographic characteristics (2). Other studies from Zambia have documented full immunization coverage estimates below national targets and have identified determinants such as child age, parity, marital status and maternal education as important predictors of whether children complete the recommended vaccination schedule (5).

North-Western Province is a largely rural region with expanding mining activities and hard-to-reach populations, including communities affected by seasonal flooding and limited road access. Provincial immunization performance has historically been broadly comparable to national averages, yet within-province variation is substantial. Earlier programme reports showed DPT3 coverage around the mid-80 percent range in North-Western, but more recent analyses and campaign data have revealed districts such as Chavuma with measles–rubella campaign coverage below 60%, attributed to geographic isolation and access barriers. At the same time, national initiatives like targeted “total control of child immunization” projects emphasise that, despite high coverage for some antigens, overall immunization coverage still leaves many children under-or unvaccinated, signalling persistent gaps that require locally tailored strategies (1,6).

Understanding temporal trends, and patterns of childhood immunization coverage among under-five children in North-Western Province is therefore critical for provincial planning and to inform Zambia’s broader efforts to reach zero-dose and under-immunized children. Analyses based on routine health information system data can show whether coverage for key antigens has improved, stagnated or declined between 2021 and 2025, and can reveal intra-provincial differences by district, facility catchment area or urban–rural setting. At the same time, examining determinants such as child age group, sex, maternal and household characteristics and service-related factors can guide targeted outreach, defaulter tracing and communication strategies (3–5).

In this context, the present retrospective analysis uses routine health information system data to describe trends and patterns in childhood immunization coverage among under-five children in North-Western Province over a 5 year period and to identify determinants associated with variations in coverage. By focusing on a single province and leveraging longitudinal facility-reported data, the study aims to generate evidence that is directly applicable to provincial micro-planning and supportive supervision, while also contributing to the national discussion on how to sustain high coverage and close equity gaps in childhood immunization.

## 2.0 Methods

This study employed a retrospective ecological design to examine spatial and temporal patterns in routine childhood immunization coverage across the districts of North-Western Province, Zambia, from 2021 to 2025. The analysis used routinely reported district-level administrative immunization data to assess temporal changes in coverage, quantify spatial inequality, and identify geographic patterns of clustering.

North-Western Province is one of Zambia’s ten provinces and comprises geographically diverse districts with predominantly rural populations. The province includes communities with varying levels of access to health services, with some areas characterised by long travel distances, difficult terrain, and seasonal constraints on accessibility. These geographic and health-service characteristics provide an important context for examining variation in routine childhood immunization coverage across districts.

The province comprised 11 districts during the study period: Chavuma, Ikelenge, Kabompo, Kalumbila, Kasempa, Mufumbwe, Mushindamo, Mwinilunga, Solwezi, Zambezi, and Zambezi.

### 2.2 Study period and population

The study covered the period from 1 January 2021 to 31 December 2025. The study population comprised children eligible for routine childhood immunization services and residing in the districts of North-Western Province during the study period. The district-year was the unit of analysis, with immunization coverage aggregated at the district level for each calendar year. Accordingly, the analysis focused on between-district geographic variation and within-district changes in immunization coverage over time, rather than individual-level vaccination status. Because the study used aggregated routine health information system data, no individual-level vaccination records were analysed.

### 2.3 Data source and data extraction

Routine childhood immunization data were obtained from the District-level immunization data were extracted from the Zambia Ministry of Health District Health Information Software 2 (DHIS2) routine health information system and compiled at district level for the five-year study period. Data included annual administrative immunization coverage indicators for routine childhood vaccines, with particular emphasis on indicators used to assess programme performance and completion of key childhood vaccination schedules. For each district and year, the available immunization coverage indicators were extracted and organised into a structured analytical dataset. The dataset was reviewed for completeness and internal consistency before analysis. Data checks included identification of missing observations, implausible values and administrative coverage estimates exceeding 100%. Where necessary, reported values were reviewed against the original routine health information records before inclusion in the analysis.

The final dataset was structured to contain a unique record for each district-year observation, together with the corresponding immunization coverage indicator(s).

Geographic boundaries for the districts were obtained from the corresponding administrative spatial dataset and linked to the immunization data using district names or unique geographic identifiers.

### 2.4 Study Variables

The **primary outcome** was district-level routine childhood immunization coverage, expressed as a percentage for each selected immunization indicator and calendar year.

The main spatial, temporal, and outcome variables were defined as follows:

- **District:** the geographic unit of analysis used to assess spatial variation and conduct spatial autocorrelation and clustering analyses.
- **Year:** calendar year from 2021 to 2025, representing the temporal dimension of the analysis.
- **Immunization coverage:** annual administrative immunization coverage (%) for the selected routine childhood immunization indicators, calculated at district level.
- **Spatial inequality:** the degree of variation in immunization coverage across districts within each year, assessed using the range, interquartile range (IQR), and coefficient of variation (CV).
- **Temporal change:** the absolute change in district-level immunization coverage between 2021 and 2025, calculated as coverage in 2025 minus coverage in 2021.

For temporal comparisons, a positive difference indicated an increase in coverage, whereas a negative difference indicated a decrease. The magnitude of the difference represented the absolute change in percentage points (pp) between the beginning and end of the study period.

Where multiple immunization indicators were analysed, each indicator was treated as a separate outcome for descriptive and spatial analyses.

### 2.5 Descriptive analysis

Descriptive statistics were used to summarise the distribution of immunization coverage across districts and years. Annual district-level coverage was presented using summary measures including the minimum, maximum, median, and measures of dispersion, where appropriate. Choropleth maps were used to visualise the geographic distribution of immunization coverage for each study year and to identify apparent differences in coverage between districts. Temporal patterns were assessed by comparing district-level coverage across the five-year study period. Changes between 2021 and 2025 were quantified using the absolute difference in coverage:

### 2.6.0 Temporal change = Coverage (2025) – Coverage (2021)

This measure was used to identify districts experiencing increases or decreases in immunization coverage over the study period.

#### 2.6.1 Assessment of spatial inequality

Spatial inequality in immunization coverage was assessed using three complementary measures; range, interquartile range (IQR), and coefficient of variation (CV).

The range was calculated as the difference between the highest and lowest district-level coverage;

#### 2.6.2 Range = Maximum coverage - Minimum coverage

The IQR was calculated as the difference between the 75th and 25th percentiles;

**IQR = Q(3)−Q(1)**

The coefficient of variation was calculated as;

**CV = (Standard deviation / Mean) × 100**

These measures were used to quantify the extent of variation in immunization coverage between districts and to complement the spatial autocorrelation analyses.

### 2.7.0 Spatial analysis

Spatial analysis was undertaken to determine whether immunization coverage was geographically clustered rather than randomly distributed across districts. District-level coverage was analysed separately for the study years, allowing spatial patterns to be examined over time. A spatial weights matrix was constructed using first-order Queen contiguity, whereby two districts were considered neighbours if they shared either a common boundary or a common vertex. The resulting spatial weights matrix was row-standardized before conducting spatial statistical analyses. Queen contiguity was selected because district-level immunization performance may plausibly be influenced by geographically adjacent districts through shared borders, population movement and service-use patterns.

#### 2.7.1 Global spatial autocorrelation

Global spatial autocorrelation was assessed using Global Moran’s I to determine whether districts with similar or dissimilar levels of immunization coverage were spatially clustered across the province. Moran’s I values greater than zero indicated positive spatial autocorrelation, suggesting clustering of similar coverage values, whereas values below zero indicated negative spatial autocorrelation, suggesting a dispersed pattern.

For each year, the observed statistic was compared with a reference distribution generated from 999 random permutations of the observed immunization coverage values across the 11 district. A two-sided significance level of α = 0.05 was used to determine whether the observed spatial autocorrelation differed significantly from that expected under spatial randomness.

#### 2.7.2 Local spatial autocorrelation

Local spatial patterns were examined using Local Moran’s I, also referred to as Local Indicators of Spatial Association (LISA). LISA was used to identify individual districts contributing to local spatial clustering and to classify districts according to their association with neighbouring districts. Significant local clusters were classified into the conventional spatial association categories of;

- High–High (HH): a district with high immunization coverage surrounded by districts with similarly high coverage;
- Low–Low (LL): a district with low coverage surrounded by districts with similarly low coverage;
- High–Low (HL): a high-coverage district surrounded predominantly by low-coverage districts; and
- Low–High (LH): a low-coverage district surrounded predominantly by high-coverage districts.

#### 2.7.3 Getis-Ord Gi* hotspot analysis

The Getis-Ord Gi* statistic was used to identify statistically significant geographic concentrations of high and low immunization coverage. The analysis identified districts forming statistically significant hotspots, representing concentrations of relatively high coverage, and coldspots, representing concentrations of relatively low coverage. Hotspot significance was assessed using 999 permutations and an α level of 0.05. The resulting z-scores and significance levels were used to distinguish statistically significant hotspots and coldspots from areas without significant spatial concentration.

#### 2.7.4 Temporal-spatial comparison

Spatial analyses were repeated across the study years to examine how the geographic distribution and clustering of immunization coverage changed over time. Annual maps of coverage, LISA clusters and Getis-Ord Gi* hotspots were compared to identify districts with persistent, emerging or changing spatial patterns. Changes between 2021 and 2025 were additionally assessed using the district-level temporal difference in coverage. This approach allowed the analysis to distinguish between districts that maintained consistently high or low coverage and those that experienced substantial improvements or declines during the study period.

#### 2.7.5 Statistical significance and analytical approach

For all spatial autocorrelation and hotspot analyses, statistical significance was assessed using 999 permutations with α = 0.05. The use of a common first-order Queen contiguity spatial weights matrix and row standardisation ensured that spatial relationships were defined consistently across districts and study years. The analysis therefore combined descriptive statistics, measures of spatial inequality, global spatial autocorrelation, local spatial autocorrelation, hotspot analysis and temporal comparison to provide a comprehensive assessment of geographic and temporal variation in routine childhood immunization coverage across North-Western Province.

## 3.0 Results

The analysis examined the spatial distribution, temporal variation, and spatial dependence of routine first-year childhood immunization coverage across the 11 districts of North-Western Province from 2021 to 2025. District-level coverage was first mapped to describe geographic variation over time, followed by assessment of spatial inequality using the range, interquartile range (IQR), and coefficient of variation (CV). Global Moran’s I was then used to assess overall spatial autocorrelation, while Local Moran’s I and Getis-Ord Gi* statistics were used to identify local clusters, spatial outliers, and statistically significant hot spots and cold spots. Spatial relationships were defined using first-order Queen contiguity based on shared district boundaries.

**Table 1.** District-level first-year childhood immunization coverage across North-Western Province, Zambia, 2021–2025.

| District | 2021 | 2022 | 2023 | 2024 | 2025 |
| --- | --- | --- | --- | --- | --- |
| Chavuma | 61.32 | 32.12 | 20.73 | 52.41 | 60.00 |
| Ikelenge | 76.05 | 68.46 | 63.1 | 72.62 | 74.29 |
| Kabompo | 82.46 | 58.72 | 37.4 | 62.06 | 79.50 |
| Kalumbila | 81.29 | 47.34 | 35.44 | 55.55 | 79.68 |
| Kasempa | 82.44 | 53.76 | 42.82 | 95.67 | 108.51 |
| Manyinga | 86.85 | 57.33 | 34.04 | 62.74 | 75.32 |
| Mufumbwe | 73.91 | 62.98 | 25.07 | 82.05 | 90.38 |
| Mushindano | 62.05 | 79.90 | 60.78 | 73.02 | 75.19 |
| Mwinilunga | 73.15 | 62.16 | 50.21 | 60.52 | 72.09 |
| Solwezi | 107.86 | 37.35 | 27.86 | 42.84 | 60.02 |
| Zambezi | 85.81 | 71.89 | 64.89 | 86.11 | 65.83 |

*FIC, first-year childhood immunization coverage. Values represent the percentage of children receiving the first-year immunization dose according to routine administrative data. Coverage estimates greater than 100% indicate that the reported number of vaccinated children exceeded the estimated target population and should therefore be interpreted with caution. District boundaries are based on official administrative boundary data.*

**Table 2.**
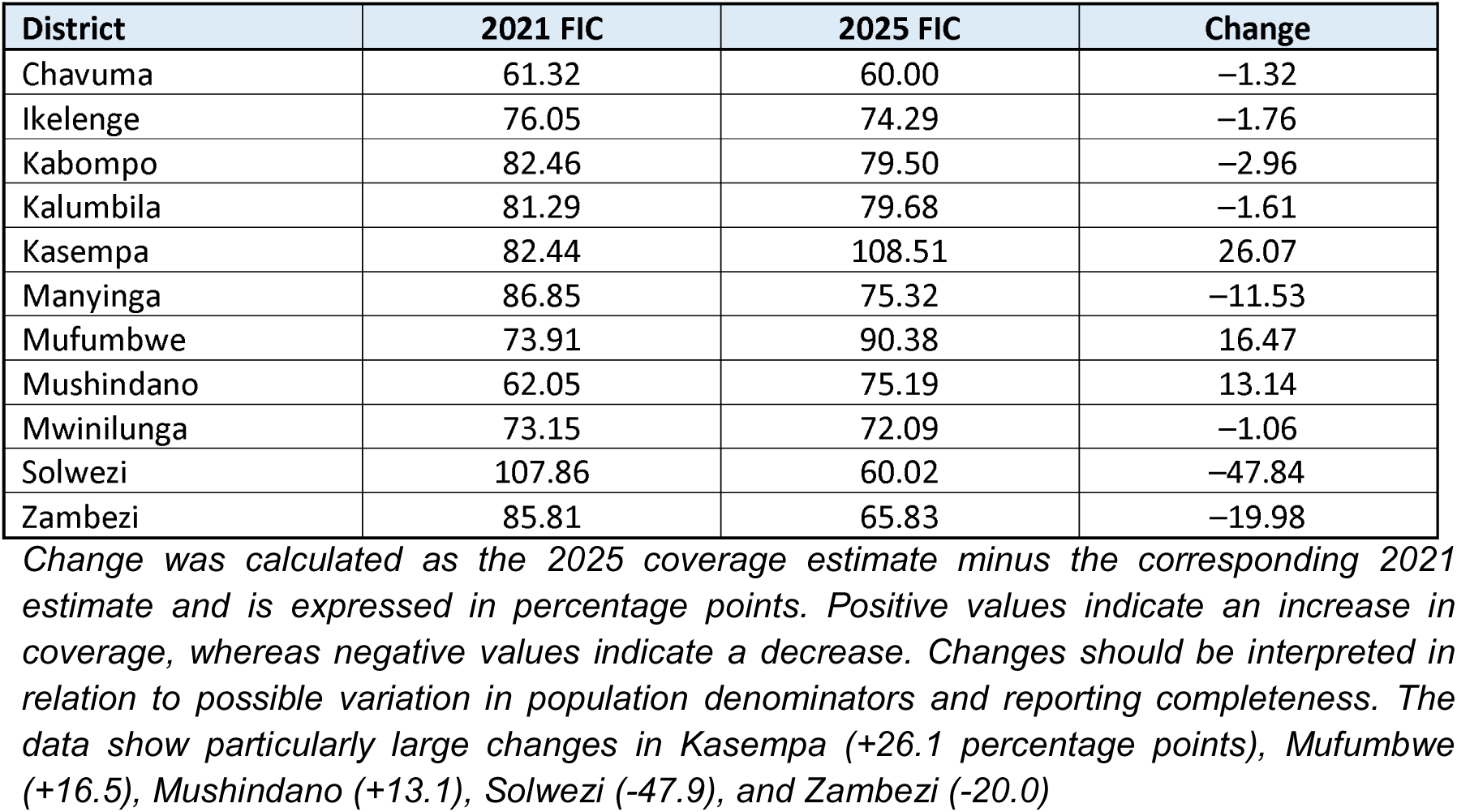
Change in first-year childhood immunization coverage between 2021 and 2025 by district.

| District | 2021 FIC | 2025 FIC | Change |
| --- | --- | --- | --- |
| Chavuma | 61.32 | 60.00 | -1.32 |
| Ikelenge | 76.05 | 74.29 | -1.76 |
| Kabompo | 82.46 | 79.50 | -2.96 |
| Kalumbila | 81.29 | 79.68 | -1.61 |
| Kasempa | 82.44 | 108.51 | 26.07 |
| Manyinga | 86.85 | 75.32 | -11.53 |
| Mufumbwe | 73.91 | 90.38 | 16.47 |
| Mushindano | 62.05 | 75.19 | 13.14 |
| Mwinilunga | 73.15 | 72.09 | -1.06 |
| Solwezi | 107.86 | 60.02 | -47.84 |
| Zambezi | 85.81 | 65.83 | -19.98 |

### 3.2.1 Location of North-Western Province within Zambia and its constituent districts

**Fig 1.**
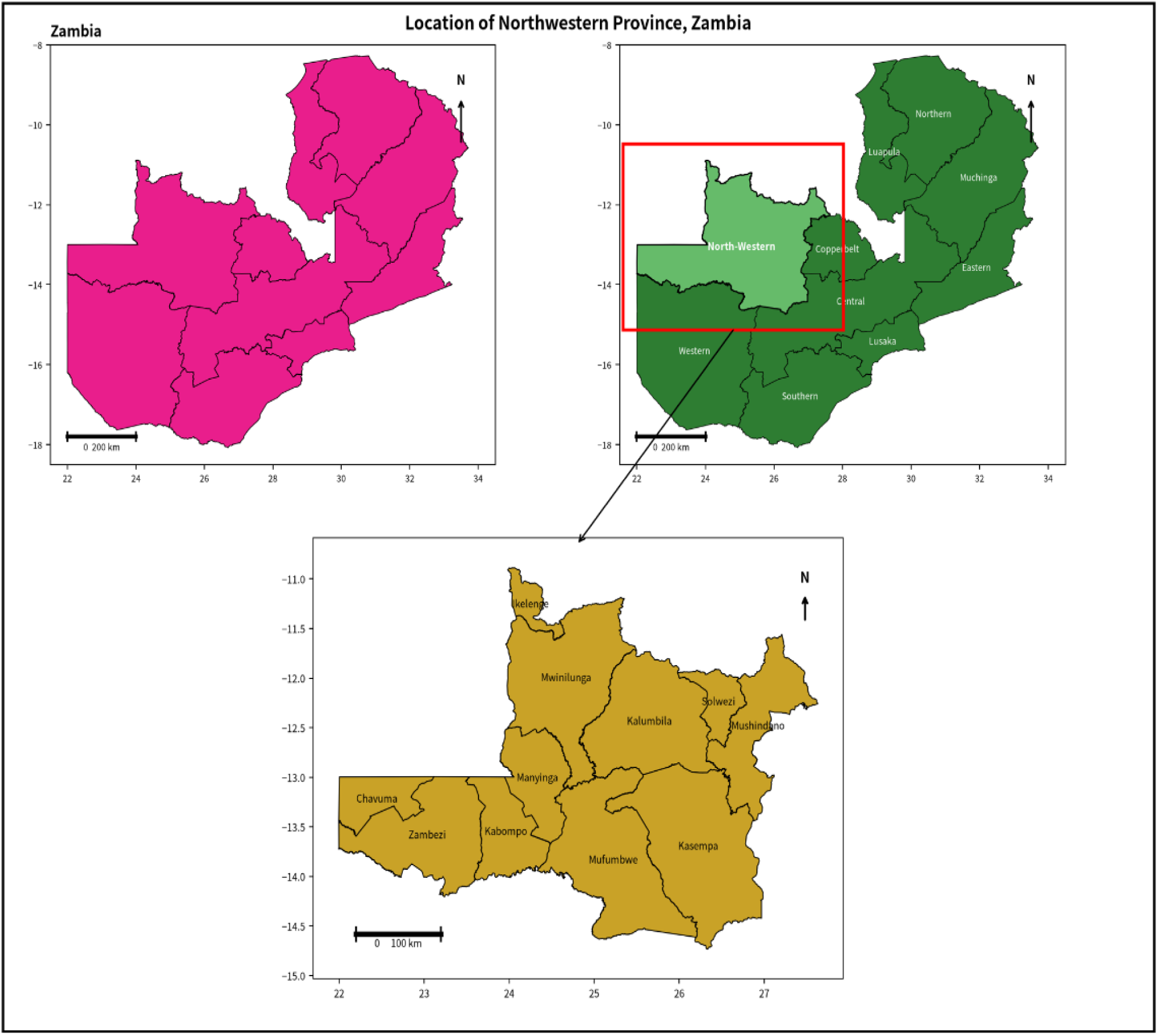
The figure shows the location of North-Western Province which is among the 10 provinces in Zambia with 11 districts with a population estimates of 1,478,574 all ages and 73,354 under ones. The province shares international borders with Angola and the Democratic Republic of the Congo (DRC).

### 3.3.2 Spatial distribution of first-year childhood immunization coverage across districts, 2021–2025

**Fig 2.**
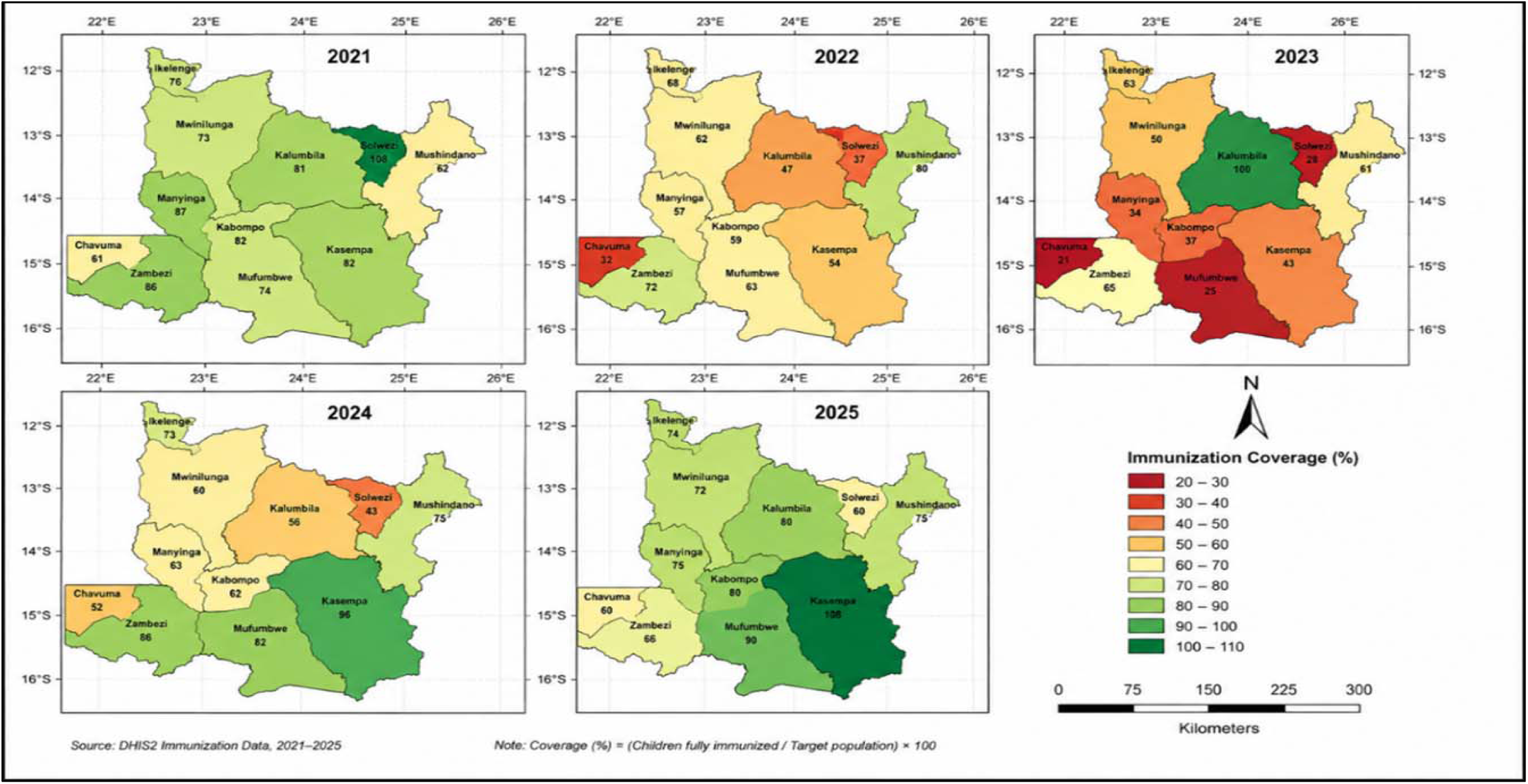
Choropleth maps show district-level FIC coverage for each year. A common classification scale was used across years to facilitate comparison of spatial patterns over time. Coverage estimates ranged from 20.7% in Chavuma in 2023 to 108.5% in Kasempa in 2025.

#### 3.4 Forest Plots – District Immunization Coverage, from 2021 to 2025 in North-Western Province, Zambia

**Fig 3.**
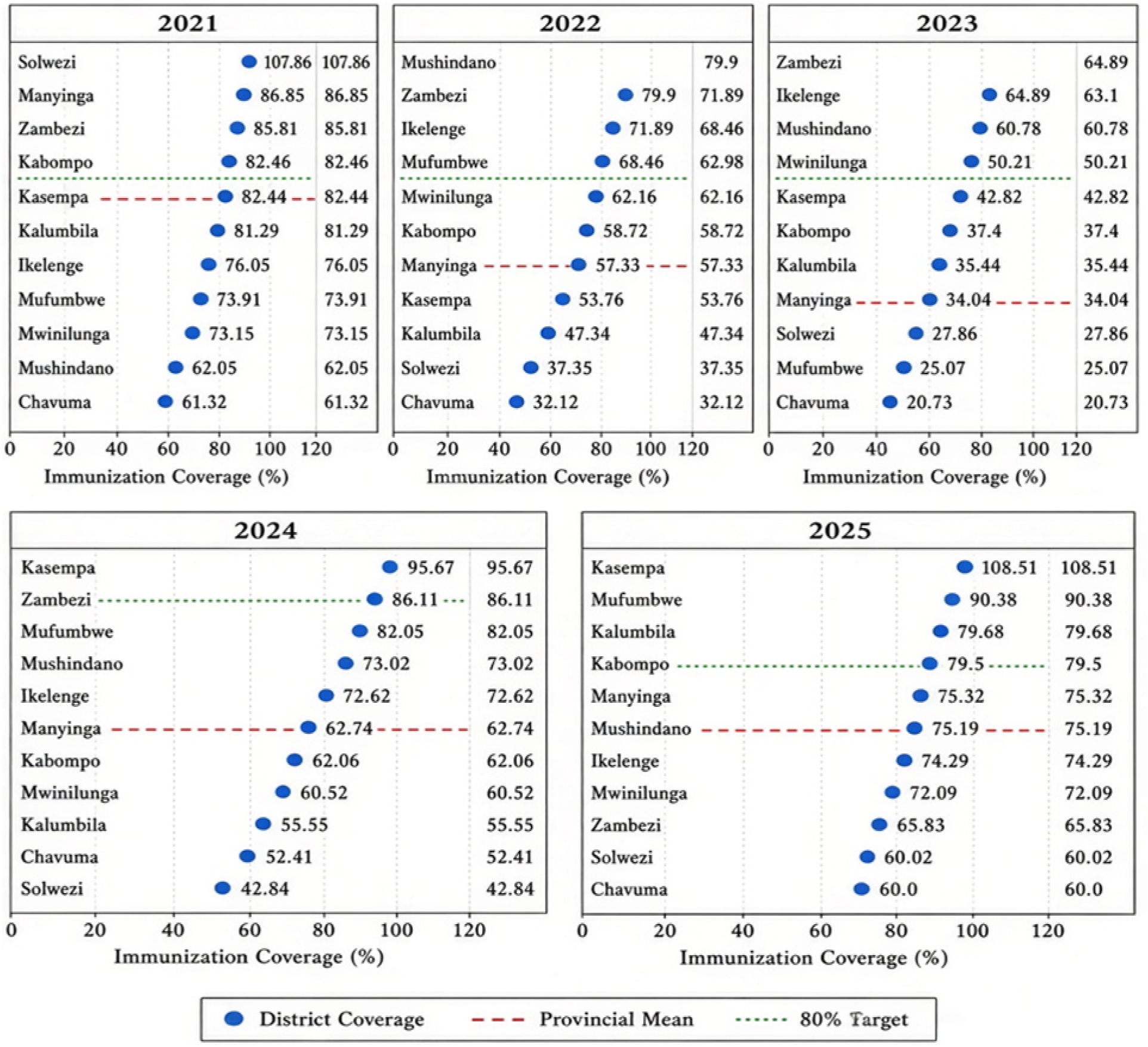
Forest plots showing district-level fully immunised coverage (%) among children under one year of age in North-Western Province, Zambia, 2021–2025. Districts are ordered from highest to lowest coverage within each year. The red dashed line represents the provincial mean coverage for that year, and the green dotted line indicates the 80% national target.

**Table 3.** Spatial variation and inequality in first-year childhood immunization coverage across districts, 2021–2025.

| Year | Mean | Min | Max | Range | IQR | CV (%) |
| --- | --- | --- | --- | --- | --- | --- |
| 2021 | 79.38 | 61.32 | 107.86 | 46.54 | 10.6 | 16.1 |
| 2022 | 57.46 | 32.12 | 79.90 | 47.78 | 15.17 | 24.9 |
| 2023 | 42.03 | 20.73 | 64.89 | 44.16 | 24.55 | 37.3 |
| 2024 | 67.78 | 42.84 | 95.67 | 52.83 | 19.50 | 23.3 |
| 2025 | 76.44 | 60.00 | 108.51 | 48.51 | 10.63 | 18.1 |

*Spatial inequality was assessed using the range, interquartile range (IQR), and coefficient of variation (CV). The range represents the difference between the maximum and minimum district-level coverage, the IQR represents the middle 50% of district values, and the CV represents relative dispersion around the mean. Higher CV values indicate greater relative variation between districts. The results show that the CV increased from 16.1% in 2021 to 37.3% in 2023, before declining to 18.1% in 2025, while the absolute range remained between 44.2 and 52.9 percentage points*

**Table 4.** Local spatial clusters and spatial outliers of first-year childhood immunization coverage, 2021, 2023 and 2025.

| Year | Moran's I | p (sim) | Interpretation ($\alpha = 0.05$) |
| --- | --- | --- | --- |
| 2021 | −0.363 | 0.055 | Not significant (near random) |
| 2022 | −0.478 | 0.021 | Significant dispersion |
| 2023 | −0.391 | 0.048 | Significant dispersion |
| 2024 | −0.053 | 0.780 | Not significant (random) |
| 2025 | 0.258 | 0.047 | Significant clustering |

*Local Indicators of Spatial Association (LISA) based on Local Moran's I were used to identify statistically significant local clusters and spatial outliers. High-High (HH) and Low-Low (LL) indicate districts with values similar to those of neighbouring districts, whereas High-Low (HL) and Low-High (LH) indicate spatial outliers. Spatial relationships were defined using row-standardized first-order Queen contiguity weights. Statistical significance was assessed using 999 permutations at $\alpha = 0.05$ . Chavuma was a significant Low-High outlier in 2023, while Kasempa and Mufumbwe formed significant High-High clusters in 2025*

#### 3.5 Local Moran’s I (LISA) Cluster Maps from 2021 to 2025 for Routine Immunization Coverage

**Fig 4.**
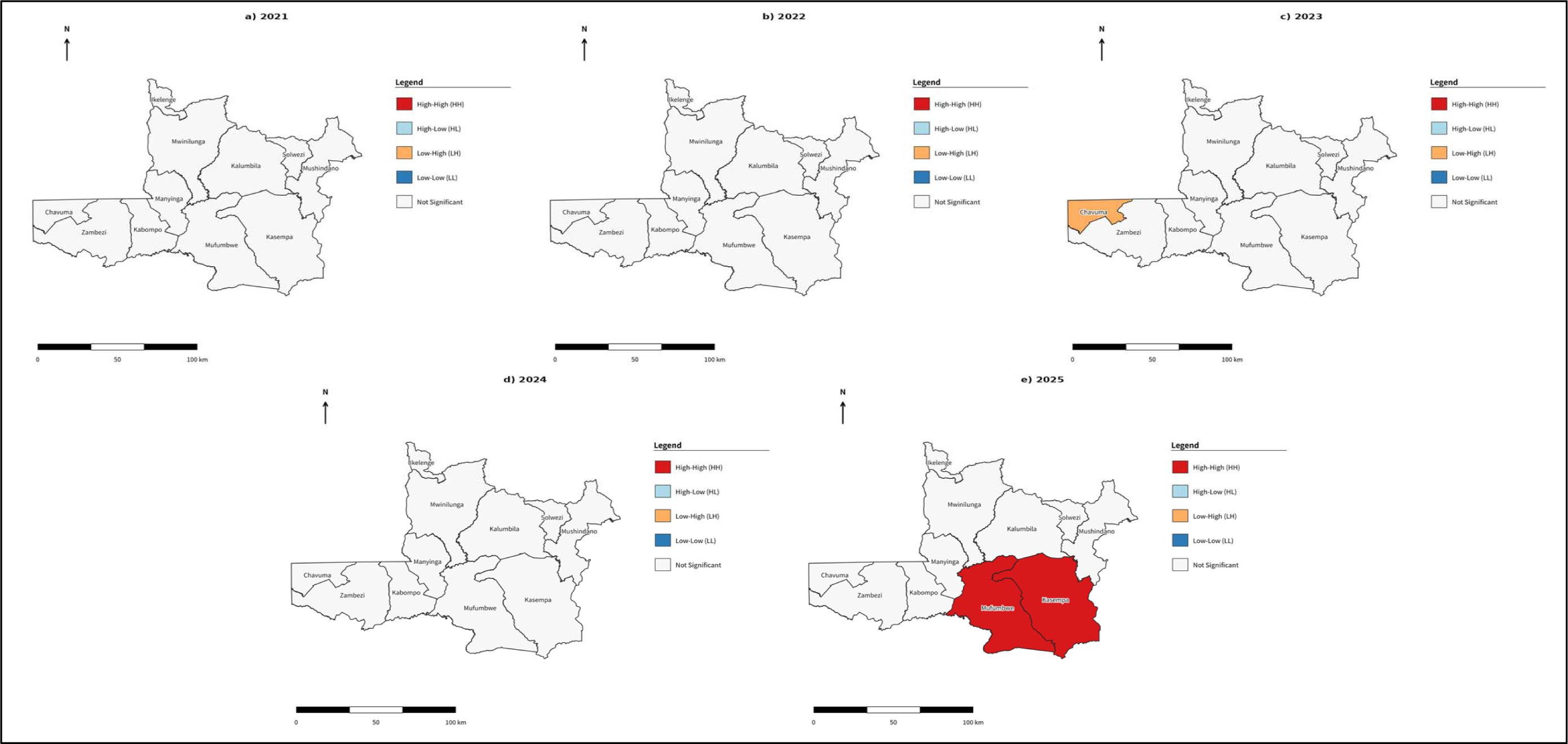
Most local statistics are non-significant. Notable results (p < 0.05); in 2023, Chavuma = significant Low-High (LH) outlier (very low coverage adjacent to higher neighbours). In 2025, Kasempa = significant High-High (HH); Mufumbwe = significant High-High (HH) while other districts show non-significant HH/HL/LH/LL patterns in most years. LISA maps for 2021, 2023 and 2025.

**Table 5.** Temporal change in first-year childhood immunization coverage (2021 and 2025)

| District | 2021 | 2025 | Change (pp) |
| --- | --- | --- | --- |
| Chavuma | 61.32 | 60.00 | -1.32 |
| Ikelenge | 76.05 | 74.29 | -1.76 |
| Kabompo | 82.46 | 79.50 | -2.96 |
| Kalumbila | 81.29 | 79.68 | -1.61 |
| Kasempa | 82.44 | 108.51 | 26.07 |
| Manyinga | 86.85 | 75.32 | -11.53 |
| Mufumbwe | 73.91 | 90.38 | 16.47 |
| Mushindano | 62.05 | 75.19 | 13.14 |
| Mwinilunga | 73.15 | 72.09 | -1.06 |
| Solwezi | 107.86 | 60.02 | -47.84 |
| Zambezi | 85.81 | 65.83 | -19.98 |

**Table 6.** Temporal trends in first-year childhood immunization coverage by district, North-Western Province, Zambia, 2021–2025.

| District | 2021 (%) | 2022 (%) | 2023 (%) | 2024 (%) | 2025 (%) | Mean (%) | Annual trend (pp/year) | Change 2021–2025 (pp) |
| --- | --- | --- | --- | --- | --- | --- | --- | --- |
| Chavuma | 61.32 | 32.12 | 20.73 | 52.41 | 60.00 | 45.32 | 1.76 | -1.32 |
| Ikelenge | 76.05 | 68.46 | 63.10 | 72.62 | 74.29 | 70.9 | 0.06 | -1.76 |
| Kabompo | 82.46 | 58.72 | 37.40 | 62.06 | 79.50 | 64.03 | -0.26 | -2.96 |
| Kalumbila | 81.29 | 47.34 | 35.44 | 55.55 | 79.68 | 59.86 | 0.50 | -1.61 |
| Kasempa | 82.44 | 53.76 | 42.82 | 95.67 | 108.51 | 76.64 | **9.40** | **26.07** |
| Manyinga | 86.85 | 57.33 | 34.04 | 62.74 | 75.32 | 63.26 | **-1.77** | **-11.53** |
| Mufumbwe | 73.91 | 62.98 | 25.07 | 82.05 | 90.38 | 66.88 | **5.20** | **16.47** |
| Mushindano | 62.05 | 79.90 | 60.78 | 73.02 | 75.19 | 70.19 | **1.94** | **13.14** |
| Mwinilunga | 73.15 | 62.16 | 50.21 | 60.52 | 72.09 | 63.63 | -0.38 | -1.06 |
| Solwezi | **107.86** | 37.35 | 27.86 | 42.84 | 60.02 | 55.19 | **-9.02** | **-47.84** |
| Zambezi | 85.81 | 71.89 | 64.89 | 86.11 | 65.83 | 74.91 | **-2.57** | **-19.98** |

#### 3.6 Getis-Ord Gi hot-spot analysis of first-year childhood immunization coverage, 2021–2025

**Fig 5.**
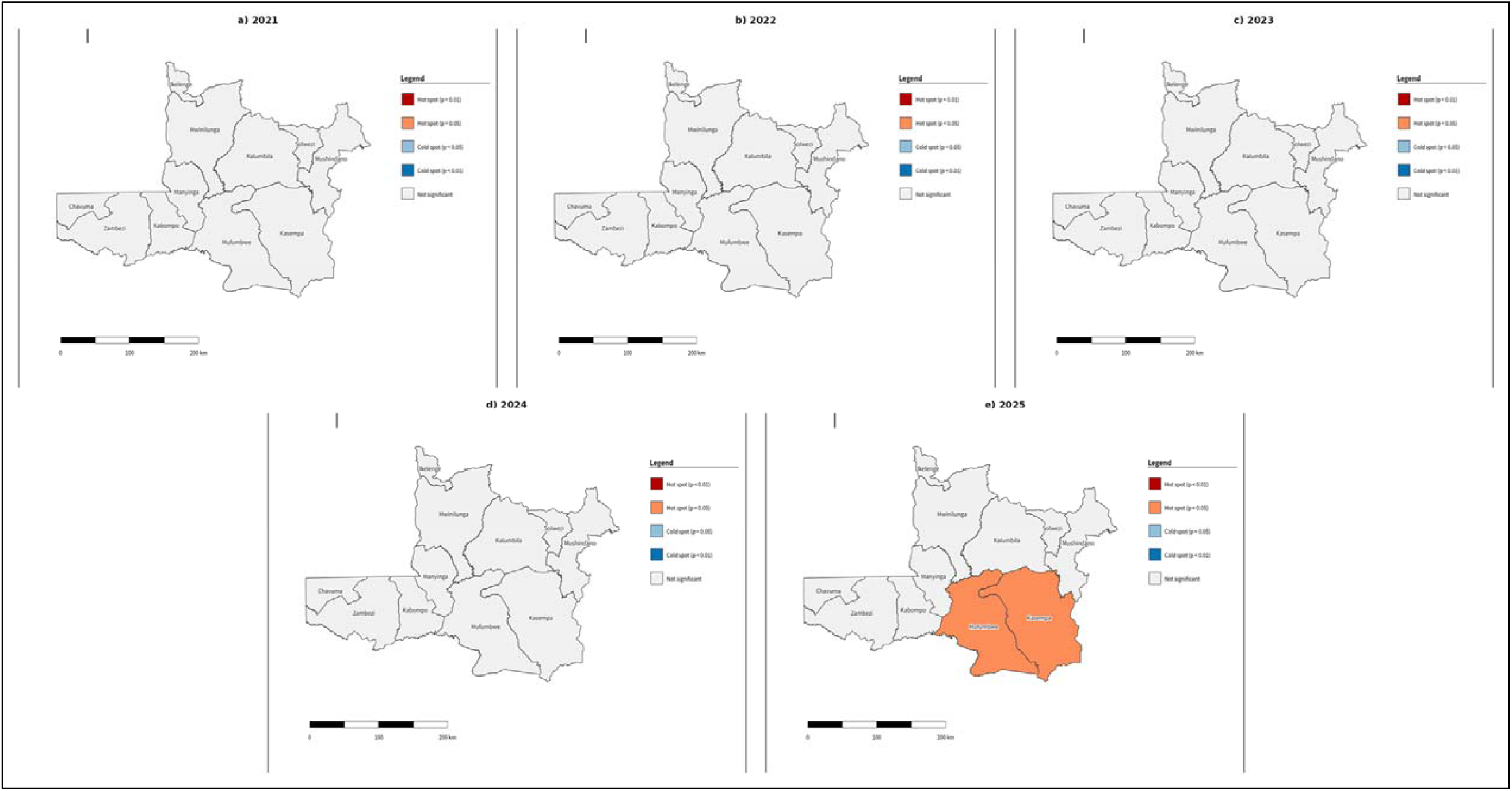
The Getis-Ord Gi* statistic was used to identify statistically significant concentrations of high and low coverage among neighbouring districts. First-order Queen contiguity spatial weights were row-standardized, and statistical significance was assessed using 999 permutations. Hot spots represent statistically significant concentrations of high coverage, whereas cold spots represent concentrations of low coverage. Statistical significance was defined as p < 0.05.

**Table 6.**
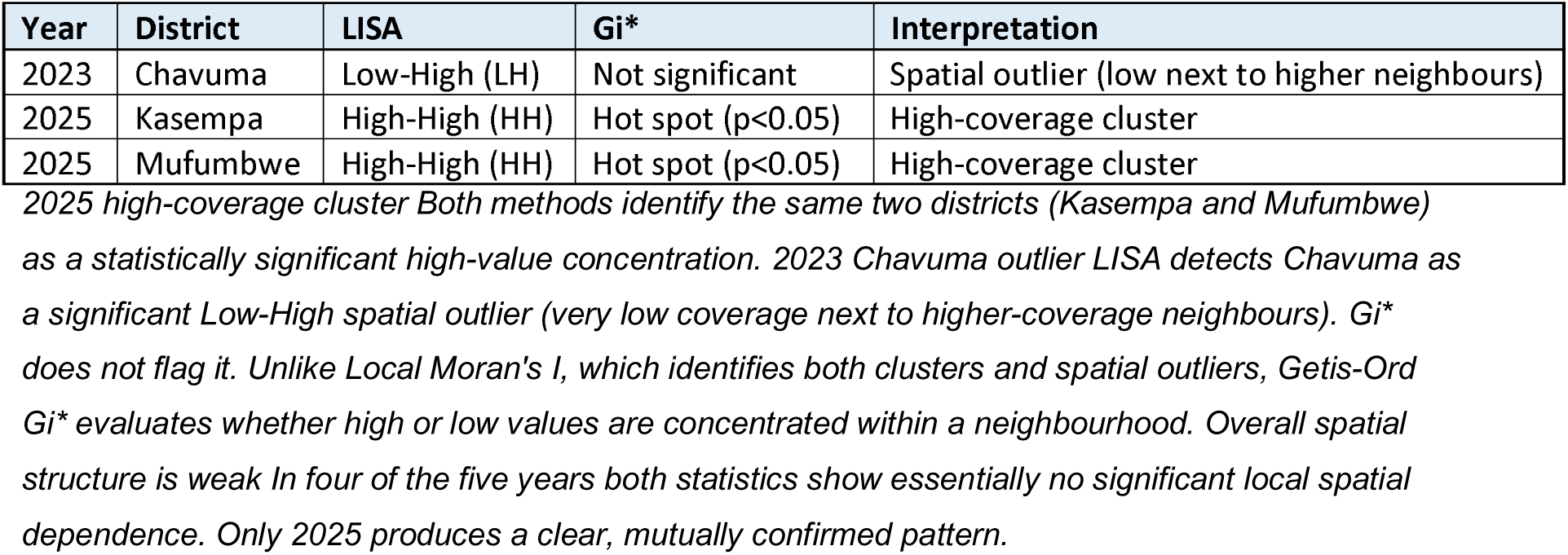
Comparison of statistically significant spatial patterns identified by Local Moran’s I and Getis-Ord Gi* across districts, 2021–2025.

| Year | District | LISA | Gi* | Interpretation |
| --- | --- | --- | --- | --- |
| 2023 | Chavuma | Low-High (LH) | Not significant | Spatial outlier (low next to higher neighbours) |
| 2025 | Kasempa | High-High (HH) | Hot spot ($p < 0.05$) | High-coverage cluster |
| 2025 | Mufumbwe | High-High (HH) | Hot spot ($p < 0.05$) | High-coverage cluster |

*2025 high-coverage cluster Both methods identify the same two districts (Kasempa and Mufumbwe)*
*as a statistically significant high-value concentration. 2023 Chavuma outlier LISA detects Chavuma as a significant Low-High spatial outlier (very low coverage next to higher-coverage neighbours). Gi\* does not flag it. Unlike Local Moran's I, which identifies both clusters and spatial outliers, Getis-Ord Gi\* evaluates whether high or low values are concentrated within a neighbourhood. Overall spatial structure is weak In four of the five years both statistics show essentially no significant local spatial dependence. Only 2025 produces a clear, mutually confirmed pattern.*

## 4.0 Discussion

This study demonstrates substantial temporal and spatial heterogeneity in routine childhood immunization coverage across districts of North-Western Province, Zambia, between 2021 and 2025. Three principal findings emerge; first, immunization performance declined markedly between 2021 and 2023 before recovering in 2024–2025; second, spatial inequality increased during the period of lowest overall coverage and subsequently narrowed as coverage recovered; and third, despite substantial differences between districts, there was little evidence of persistent global spatial autocorrelation, with statistically significant local spatial structures emerging only in specific years, most notably the high-coverage cluster involving Kasempa and Mufumbwe in 2025. These findings suggest that immunization performance in the province is characterized less by a persistent province-wide geographical divide than by temporally shifting pockets of relative underperformance and high performance.

### 4.1 Temporal Changes in Immunization Coverage

The most prominent finding was the pronounced decline in immunization coverage between 2021 and 2023. The provincial mean decreased from 79.4% in 2021 to 57.5% in 2022 and 42.0% in 2023, before increasing to 67.8% in 2024 and 76.4% in 2025. The 2023 nadir was accompanied by a marked increase in between-district variation, with the coefficient of variation increasing from 16.1% in 2021 to 37.3% in 2023. The temporal pattern occurred within the period of documented disruption and subsequent recovery of routine immunization services globally, although the ecological design and absence of service-level explanatory variables prevent attribution of the observed provincial decline specifically to the COVID-19 pandemic. The World Health Organization (WHO) has documented substantial disruption to routine immunization services during the pandemic, including interruptions to outreach activities, reduced demand for vaccination, transportation barriers and difficulties accessing health facilities. The African Region was particularly affected, with routine immunization coverage declining substantially during 2020 and 2021 and recovery subsequently occurring unevenly across countries and populations (7,8).

The timing of the decline observed in North-Western Province should nevertheless be interpreted cautiously. The present study is based on district-level programme data and therefore cannot establish that COVID-19 was the direct cause of the observed decline. The 2021 to 2023 pattern may reflect a combination of service disruption, changes in outreach intensity, vaccine availability, health-worker capacity, caregiver demand, population mobility and changes in administrative denominators. This distinction is important because the study does not contain information on the individual mechanisms responsible for the observed changes. Nevertheless, the temporal coincidence with the documented global disruption of routine immunization provides a plausible programme-level context for the decline. The subsequent recovery between 2023 and 2025 is also consistent with the broader international emphasis on catch-up vaccination, restoration of routine services and strengthening of primary health-care systems following the pandemic (7).

The recovery in 2024 to 2025 is encouraging but should not be interpreted as evidence that immunization performance has fully returned to a uniformly high and equitable level. WHO’s most recent global assessment indicates that, despite recovery in some indicators, routine vaccination coverage remains insufficient to place the world on track for the Immunization Agenda 2030 (IA2030) targets. In 2025, global DTP3 coverage remained approximately 85%, while 13.5 million children received no initial DTP dose. WHO and UNICEF have consequently emphasized the importance of strengthening routine systems and using subnational data to identify populations that remain unreached (9).

The recovery observed in this study is therefore best understood as a positive programme response rather than the resolution of immunization inequities. This distinction is supported by the 2025 IA2030 mid-term review, which concluded that many immunization targets remain off track and called for stronger integration with primary health care, improved data-driven decision-making and renewed attention to zero-dose and under-immunized populations (9).

### 4.2 Persistent Differences between Districts

Although the overall provincial trend was characterized by decline followed by recovery, individual districts followed markedly different trajectories. Kasempa recorded the largest improvement between 2021 and 2025 (+26.1 percentage points), followed by Mufumbwe (+16.5) and Mushindano (+13.1). In contrast, Solwezi experienced the largest reported decline (−47.9 percentage points), followed by Zambezi (−20.0) and Manyinga (−11.5).

The divergent trajectories indicate that provincial averages can obscure important district-level differences. This is consistent with the increasing emphasis by WHO and UNICEF on subnational immunization monitoring. WHO notes that national or aggregate coverage can conceal substantial inequalities within countries and that subnational monitoring is necessary to identify districts requiring focused interventions. In 2025, WHO collected subnational immunization data from 147 Member States, further highlighting the growing importance of geographically disaggregated surveillance and programme monitoring (10,11).

The improvement observed in Kasempa and Mufumbwe is particularly noteworthy because both districts subsequently formed a statistically significant high-coverage spatial cluster in 2025. However, the available results do not establish which programme interventions produced these improvements. Possible explanations such as improved outreach, better microplanning, increased health-worker performance, improved data reporting or stronger community engagement should therefore be considered hypotheses for future investigation rather than conclusions from the present analysis. Similarly, the decline in Solwezi requires particularly cautious interpretation. The results indicate that the target population or denominator changed substantially between study years, and the research appropriately identifies this as a potential explanation for the very large apparent decline. Administrative immunization coverage can exceed 100% when vaccination doses are attributed to a denominator that underestimates the population eligible for vaccination, when children from outside the reporting area receive services within the district, or when numerator and denominator populations are inconsistent. WHO likewise cautions that the quality and comparability of administrative coverage estimates depend heavily on the underlying reported data and denominators (11,12).

Consequently, the exceptionally high values observed in Solwezi in 2021 and Kasempa in 2025 should not be interpreted literally as evidence that more than 100% of eligible children were biologically immunized. Instead, they may indicate limitations in population denominators, population mobility, service utilization across district boundaries or reporting practices. This issue is especially important in geographically mobile populations and districts that attract patients from neighbouring areas.

### 4.3 Spatial Inequality Increased During the Coverage Trough

An important contribution of this study is the demonstration that spatial inequality was greatest when overall immunization performance was poorest. The coefficient of variation increased from 16.1% in 2021 to 24.9% in 2022 and 37.3% in 2023, while the interquartile range (IQR) increased from 10.6 percentage points to 24.6 percentage points over the same period. By 2025, however, the IQR had declined to 10.7 percentage points and the CV to 18.1%.

This finding suggests that deterioration in immunization performance was not spatially uniform. Some districts experienced considerably greater reductions than others, resulting in widening disparities during the 2023 trough. Conversely, recovery was accompanied by narrowing relative inequality. Such a pattern is important from an equity perspective because an improvement in the provincial mean can occur without corresponding improvement among the lowest-performing districts.

Evidence from sub-Saharan Africa supports the importance of examining immunization inequalities below the national level. An analysis of Demographic and Health Survey data from 25 sub-Saharan African countries found substantial socioeconomic inequalities in childhood vaccination, with zero-dose children disproportionately concentrated among disadvantaged populations; higher maternal education, greater household wealth, facility delivery and adequate antenatal care were associated with higher vaccination uptake (13). More recent geospatial evidence further demonstrates that these inequalities are not uniformly distributed within countries. A multicountry analysis of 21 sub-Saharan African countries identified substantial subnational heterogeneity and localized zero-dose hotspots, with zero-dose vaccination strongly associated with rural residence, low household wealth, limited maternal education, restricted information access, home delivery and inadequate antenatal care (14). Similarly, a recent spatial analysis across 33 sub-Saharan African countries identified 27 regions with higher-than-expected concentrations of zero-dose children and found consistently higher zero-dose prevalence in areas with greater proportions of mothers reporting no antenatal care (15). Together, these findings support the value of subnational spatial analysis for identifying localized immunization inequalities that may be obscured by national or provincial averages.

The present study cannot determine whether the observed district-level differences were driven by socioeconomic status, geographical accessibility, health-system capacity or caregiver-level factors because these explanatory variables were not included in the analysis. However, the observed spatial heterogeneity supports the need to investigate these determinants. IA2030 specifically emphasizes identification of underserved populations and geographically targeted interventions rather than relying exclusively on aggregate coverage (16).

### 4.4 Limited Evidence of Persistent Global Spatial Clustering

Despite substantial variation in district performance, Global Moran’s I showed no statistically significant spatial autocorrelation in 2021, 2023 or 2024. The statistics for 2021 to 2024 were predominantly negative, although the 2022 estimate approached conventional significance (I = −0.438, p = 0.066). In 2025, Moran’s I became positive (I = 0.256, p = 0.052), indicating a shift towards weak positive spatial dependence that was close to, but did not reach, the conventional 0.05 significance threshold.

This finding is important because it indicates that districts with similar immunization performance were not consistently geographically clustered throughout the study period. Rather than demonstrating a stable province-wide spatial structure, the results suggest that the geographical organization of immunization performance changed over time. The early tendency towards negative spatial dependence is compatible with a pattern in which relatively high-and low-performing districts occur adjacent to one another, whereas the positive statistic in 2025 suggests emerging geographical concentration of similar performance.

### 4.5 Identification of a High-Coverage Cluster in 2025

The strongest spatial finding was the emergence of a statistically significant high-coverage cluster involving Kasempa and Mufumbwe in 2025. Both districts were classified as High-High clusters using Local Moran’s I and as significant hot spots using the Getis-Ord Gi* statistic. The agreement between the two approaches strengthens confidence that the observed 2025 concentration represents a meaningful local spatial pattern rather than an artefact of one clustering statistic. This pattern is noteworthy because neither LISA nor Gi* identified significant local clusters during 2021, 2022 or 2024. Thus, the 2025 cluster appears to represent an emerging spatial concentration rather than a persistent high-performing area. This temporal emergence is consistent with the observed improvement in both districts; Kasempa increased from 82.4% in 2021 to 108.5% in 2025, while Mufumbwe increased from 73.9% to 90.4%.

Comparable studies in sub-Saharan Africa demonstrate that immunization coverage can form geographically localized clusters even when national or regional averages appear relatively satisfactory. Brownwright and colleagues, using spatial analysis of measles vaccination coverage across 10 sub-Saharan African countries, identified 477 low-coverage spatial clusters, including clusters in Zambia and Malawi, and demonstrated that localized vaccination patterns could differ substantially from national coverage levels (17). More recent evidence has reinforced this finding. A 2026 multicountry analysis of 21 sub-Saharan African countries identified substantial within-country spatial heterogeneity and localized zero-dose hotspots, with most countries demonstrating significant subnational clustering (14). Similarly, an analysis of 33 sub-Saharan African countries identified 27 regions with higher-than-expected concentrations of zero-dose children, while a separate 2025 study found significant spatial clustering of incomplete childhood immunization and geographically distinct hotspot regions (15). Taken together, these findings support the interpretation that aggregate coverage measures can mask important geographic inequalities and that spatially targeted approaches may be necessary to identify and address localized immunization gaps

The emergence of Kasempa and Mufumbwe as a high-coverage cluster therefore has practical implications. Rather than treating these districts simply as areas that have achieved higher coverage, programme managers could examine the service-delivery practices, outreach strategies, community engagement mechanisms, data-management practices and resource allocation patterns operating in these districts. If specific successful practices can be identified and verified, they could potentially provide transferable lessons for districts with persistent or recurrent performance gaps.

### 4.6 Chavuma as a Spatial Outlier in 2023

In contrast to the 2025 high-coverage cluster, Chavuma was identified as a statistically significant Low-High outlier in 2023. This means that Chavuma had unusually low coverage relative to its neighbouring districts rather than forming part of a geographically contiguous cluster of low-performing districts. The distinction between a low-low cluster and a low-high outlier is programmatically important. A Low-High outlier may indicate a district-specific problem occurring within a broader area where neighbouring districts are performing relatively better. Therefore, interventions based solely on regional targeting may be less appropriate than a more focused assessment of Chavuma’s local circumstances. Potential explanations could include differences in service accessibility, outreach implementation, health-worker availability, vaccine logistics, population mobility, reporting completeness or local demand.

The finding also demonstrates the complementary value of LISA and Getis-Ord Gi*. Gi* did not classify Chavuma as a significant cold spot because Gi* identifies concentrations of similarly high or low values, whereas LISA can identify a single district whose value differs significantly from that of its neighbours. Using both methods therefore provides a more nuanced interpretation of the spatial structure than relying on either statistic alone.

### 4.7 Implications for Immunization Programme Planning

The findings have several implications for routine immunization planning in North-Western Province. First, programme monitoring should move beyond provincial averages and routinely incorporate district-level spatial analysis. WHO emphasizes that national or aggregate immunization coverage can conceal substantial inequalities in coverage and access within countries and that subnational monitoring can identify these disparities and inform focused interventions (18). The use of district-level maps and spatial statistics could therefore help provincial and district health authorities identify persistent low-coverage areas, monitor emerging high-and low-coverage clusters, and prioritize geographically targeted programme responses. This approach is consistent with the Immunization Agenda 2030, whose 2025 Mid-Term Review calls for stronger data-driven decision-making and more focused efforts to address persistent gaps in coverage and equity (9).

Second, districts should not necessarily receive identical interventions. The 2023 Chavuma Low-High pattern and the 2025 Kasempa-Mufumbwe High-High pattern demonstrate that geographically adjacent districts can exhibit markedly different coverage patterns and programme performance. These findings support an equity-oriented approach in which interventions are tailored to local performance, population needs and the barriers affecting vaccination uptake rather than applying uniform strategies across districts. This approach is consistent with the Immunization Agenda 2030 (IA2030), which calls for locally tailored, evidence-based and people-centred strategies to reach poorly served populations and for the use of subnational data to identify and track zero-dose and under-immunized populations (16). WHO further recommends moving beyond one-size-fits-all approaches by identifying where immunization inequities occur, determining their underlying barriers, implementing tailored responses and subsequently evaluating their effectiveness (19).

Third, strengthening the quality of target-population denominators and administrative reporting should be considered a central component of routine immunization improvement. The presence of coverage estimates exceeding 100% in several district-years raises concerns about the accuracy of the underlying numerator and denominator and limits the extent to which observed changes can be interpreted as true changes in vaccination uptake. WHO identifies coverage estimates substantially above 100% and marked year-to-year fluctuations as potential indicators of inaccurate target-population estimates (20). WHO also notes that administrative coverage can be biased by inaccurate numerators or denominators, with denominator inaccuracies arising from factors including population movement, inaccurate census estimates or projections, and the use of multiple denominator sources (18). Accordingly, routine reconciliation of target-population estimates, vaccination registers, facility reports and population projections, together with systematic assessment of reporting completeness and accuracy, could improve the validity and comparability of future district-level spatial analyses (21).

Finally, the absence of statistically significant cold spots indicates that the analysis did not detect geographically concentrated low-coverage values under the specified spatial weights and significance threshold; it does not imply that all districts had adequate coverage. The significant Low–High outlier identified in 2023 illustrates this distinction, as a district may exhibit relatively low coverage despite being surrounded by higher-performing neighbours without forming a conventional Low–Low cluster. Comparable spatial analyses in sub-Saharan Africa have similarly identified Low–High and High–Low outliers alongside conventional High–High and Low–Low clusters, demonstrating that immunization inequalities can be spatially heterogeneous and may not be fully captured by conventional hotspot or cold-spot analysis alone (15). Recent multicountry evidence further shows that zero-dose vaccination is associated with rural residence, socioeconomic disadvantage, limited maternal education, inadequate antenatal care, home delivery and perceived difficulties accessing health facilities (22). Consequently, future analyses should integrate routine immunization coverage with measures of geographical accessibility, socioeconomic deprivation, health-service availability, outreach intensity, vaccine availability and maternal-health service utilization to better identify the mechanisms underlying observed spatial patterns and inform appropriately targeted interventions.

### 4.8 Strengths and Limitations

A major strength of this study is the use of five consecutive years of district-level routine immunization data, which enabled assessment of both temporal changes and spatial variation over time. The application of complementary spatial methods—including choropleth mapping, measures of spatial inequality, Global Moran’s I, Local Moran’s I (LISA), and Getis-Ord Gi*—provided a multidimensional assessment of the geographic distribution and evolution of immunization coverage. The identification of the same 2025 high-coverage pattern involving Kasempa and Mufumbwe by both LISA and Getis-Ord Gi* provides convergent evidence for the presence of a localised high-coverage pattern.

Several limitations should be considered when interpreting the findings. First, the study relied on routine administrative immunization data, which may be affected by incomplete reporting, reporting inconsistencies, inaccurate target-population denominators, and differences in data quality or reporting practices between districts. The observation of coverage estimates exceeding 100% illustrates the potential influence of denominator or reporting-related issues and warrants cautious interpretation of absolute coverage levels.

Second, the ecological and district-level design precludes inference at the individual level. The observed geographic patterns cannot be interpreted as evidence that individual children with particular characteristics were more or less likely to be immunized. Similarly, the study design does not permit causal conclusions regarding factors underlying differences in immunization coverage between districts.

Third, spatial relationships were defined using first-order Queen contiguity, whereby districts were considered neighbours when they shared a common boundary or vertex. This representation may not fully capture functional spatial relationships arising from road networks, population mobility, health-service utilisation, referral patterns, or physical access to health facilities. Consequently, the identified spatial patterns may differ if alternative spatial weights matrices are used.

Fourth, the relatively small number of spatial units (11 districts) limits statistical power and may affect the stability and sensitivity of global and local spatial statistics. Local cluster and outlier identification may be particularly sensitive to the configuration of the spatial weights matrix. Therefore, isolated statistically significant local patterns should be interpreted cautiously and in conjunction with the observed coverage values and geographic context.

Finally, the analysis was primarily descriptive and spatial in nature and was designed to identify geographic and temporal patterns rather than explain their underlying causes. Factors such as health-service accessibility, vaccine supply, staffing, population mobility, socioeconomic conditions, and differences in programme implementation were not directly incorporated into the spatial analysis. Further analytical studies incorporating these factors are needed to determine the drivers of observed district-level inequalities and spatial clustering.

## 5.0 Conclusion

Routine childhood immunization coverage in North-Western Province varied substantially across districts and over time between 2021 and 2025, with the greatest spatial inequality observed in 2023 followed by a reduction in inequality in 2024 and 2025. Global spatial autocorrelation was generally weak and non-significant, suggesting that immunization coverage did not follow a consistent province-wide spatial pattern during the study period. However, localized spatial structures were evident in specific years, including the Low-High spatial outlier in Chavuma in 2023 and the statistically significant High-High cluster and hotspots involving Kasempa and Mufumbwe in 2025.

These findings demonstrate the importance of moving beyond provincial or aggregate coverage estimates towards continuous, district-level monitoring of temporal trends, spatial inequality, and local clustering. Routine programme monitoring should prioritize districts experiencing sustained or marked declines in coverage, emerging low-coverage patterns, or substantial departures from neighbouring districts. Strengthening the accuracy and consistency of target-population denominators is also essential, particularly where coverage estimates exceed 100%, as denominator inaccuracies may obscure the true magnitude and distribution of immunization gaps.

The findings further support the use of spatially informed micro-planning and targeted outreach to identify and address emerging programme gaps before they become persistent. However, spatial patterns alone cannot explain the underlying causes of variation. Further research should therefore investigate the health-system, geographic, demographic, socioeconomic, and service-access factors associated with district-level differences in immunization coverage. Understanding these contextual factors, alongside lessons from higher-performing districts, may support the development of interventions that are tailored to local needs.

Overall, integrating spatial and temporal analysis into routine immunization surveillance can strengthen the identification of underserved populations and improve the targeting of programme resources. For North-Western Province, strengthening district-level surveillance and using these data to guide context-specific micro-planning may provide a practical pathway towards more equitable and sustained childhood immunization coverage.

## 5.1 Declaration

## 5.2 Ethics approval and consent to participate

## 5.3 Ethical Considerations

This study used routinely collected, aggregated district-level health information and did not involve direct contact or interaction with individual participants. The analytical dataset contained no personally identifiable information or individual-level health records. Access to and use of the routine health information data were authorised by the relevant health authorities and conducted in accordance with applicable institutional and national requirements.

## 5.4 Consent for publication

Not applicable.

## 5.7 Availability of data and materials

The datasets generated and/or analyzed during the current study are not publicly available due to restrictions from the Ministry of Health but are available from the corresponding author on reasonable request.

## 5.9.0 Authors’ contributions

MCM conceived and designed the study, coordinated data acquisition and management, performed the statistical and spatial analyses, interpreted the findings, and drafted the initial manuscript. FN contributed to the study design, data acquisition and management, interpretation of the immunization coverage findings, and critical revision of the manuscript. EK contributed to data management, assisted with interpretation of the study findings, and critically reviewed and revised the manuscript. MC provided technical guidance on the study design and analytical approach, contributed to interpretation of the findings, and critically revised the manuscript for important intellectual content. CH contributed to the literature review, data entry and management, interpretation of findings, and editing of the manuscript, including final formatting. All authors critically reviewed the manuscript, read and approved the final version, and agreed to be accountable for all aspects of the work

### 5.9.1 Authors’ information

Marshall Chitalu Mubanga is a lecturer and researcher in public health with a focus on infectious disease epidemiology and biostatistics. Martin Chakulya is a lecturer in pathology and microbiology with expertise in infectious disease diagnostics. Chipego Hajamba is a lecturer in nursing sciences with a focus on maternal and child health. Eustarckio Kazonga is a lecturer in biostatistics and epidemiology, and researcher in public health. Friday Nkhoma is a Manager at CHAZ under his portofolio is the EPI and CSO Cordination for demand generation for Immunisation.

## Data Availability

All data produced in the present study are available upon reasonable request to the authors

## 5.8 Acknowledgements

We thank the Ministry of Health for access to Provincial Immunization coverages from the DHIS2 system and their support in data abstraction.

